# Polygenic risk scores predict diabetic complications and their response to therapy

**DOI:** 10.1101/19010785

**Authors:** J. Tremblay, M. Haloui, F. Harvey, R. Tahir, F.-C. Marois-Blanchet, C. Long, R. Attaoua, P. Simon, L. Santucci, C. Hizel, J. Chalmers, M. Marre, S. Harrap, R. Cifkova, A. Krajcoviechova, D. Matthews, B. Williams, N. Poulter, S. Zoungas, S. Colagiuri, G. Mancia, D.E. Grobbee, A. Rodgers, L. Liu, M. Agbessi, V. Bruat, M-J. Favé, M. Harwood, P. Awadalla, M. Woodward, P. Hamet

## Abstract

Type 2 diabetes increases the risk of cardiovascular and renal complications, but early risk prediction can lead to timely intervention and better outcomes. Through summary statistics of meta-analyses of published genome-wide association studies performed in over 1.2 million of individuals, we combined 9 PRS gathering genomic variants associated to cardiovascular and renal diseases and their key risk factors into one logistic regression model, to predict micro- and macrovascular endpoints of diabetes. Its clinical utility in predicting complications of diabetes was tested in 4098 participants with diabetes of the ADVANCE trial followed during a period of 10 years and replicated it in three independent non-trial cohorts. The prediction model adjusted for ethnicity, sex, age at onset and diabetes duration, identified the top 30% of ADVANCE participants at 3.1-fold increased risk of major micro- and macrovascular events (p=6.3×10^−21^ and p=9.6×10^−31^, respectively) and at 4.4-fold (p=6.8×10^−33^) increased risk of cardiovascular death compared to the remainder of T2D subjects. While in ADVANCE overall, combined intensive therapy of blood pressure and glycaemia decreased cardiovascular mortality by 24%, the prediction model identified a high-risk group in whom this therapy decreased mortality by 47%, and a low risk group in whom the therapy had no discernable effect. Patients with high PRS had the greatest absolute risk reduction with a number needed to treat of 12 to prevent one cardiovascular death over 5 years. This novel polygenic prediction model identified people with diabetes at low and high risk of complications and improved targeting those at greater benefit from intensive therapy while avoiding unnecessary intensification in low-risk subjects.

## Introduction

Diabetes increases the risk of serious and costly cardiovascular and renal complications^1,2^. Prediction of risk prior to development of these complications is crucial to enable targeting individuals that could benefit from an early intervention^3^. Current clinical risk prediction models are only applicable once clinical signs are present. Genetic information, with which one is born, offers a way to make early detection of risk. Genome-wide association studies (GWAS) identified multiple common variants associated with T2D^4-7^, renal^8,9^, cardiovascular diseases^10^, and hypertension^11^. Individually, these genetic variants account for only a small effect size but the combination of hundreds or thousands of them into polygenic risk scores (PRS) or genome-wide polygenic scores (GPS) was recently introduced to predict individual risk of diseases^12-15^ including type 2 diabetes^16^. Recent studies suggest that combining multi-PRS of related traits into a joint model could optimize its prediction performance^17,18^. Because of their common risk factors, overlap in terms of pathogenetic mechanisms and correlations among them, we combined 9 PRS gathering genomic variants associated to cardiovascular and renal diseases and their key risk factors into one logistic regression model, to predict micro- and macrovascular endpoints of diabetes^19-23^. We selected SNPs within loci associated to 26 risk factors and outcomes of T2D obtained from summary statistics data of meta-analyses of genome-wide association studies (GWASs) validated in 1.2 million of participants of European descent and grouped them into 9 PRS that were then included as variables in the logistic regression model. The prediction performance of the polygenic model was assessed by c-statistics using a target sample composed of 4098 genotyped participants of European descent of the ADVANCE trial,^24,25^ extended to its post-trial follow-up, ADVANCE-ON^26^ for a total of nearly 10 years of observation. The predictors retained in the model were the nine PRS, the first principal component (PC1)^25^ of ancestry, sex, age at diagnosis, and diabetes duration. This parsimonious polygenic prediction model did not include any clinical or outcome data and was replicated in three independent non-trial cohorts. It was used to identify individuals who benefited most (or not) from the intensive therapy administered in ADVANCE.^27,28^

## Methods

### Cohorts

#### Testing cohort

ADVANCE was a 2×2 factorial randomized controlled trial of blood pressure (BP) lowering (perindopril-indapamide *vs* placebo) and intensive glucose control (gliclazide MR-based intensive intervention with a target of 6.5 HbA1c *vs* standard care) in patients with T2D. A total of 11,140 participants were recruited from 215 centers in 20 countries. They were older than 55 years and diagnosed with T2D after the age of 30 years. The trial was successful in decreasing total and cardiovascular mortality by attenuation of combined microvascular and macrovascular primary outcome with blood pressure control^24^ and combined control of blood glucose and blood pressure.^28^ ADVANCE-ON was a 5-year post-trial observational extension of ADVANCE conducted in 80% of subjects^26^. Here, we studied a subset of 4098 genotyped T2D patients of Caucasian origin from ADVANCE.

#### Replication cohorts

The Czech post-MONICA study was a cross-sectional survey investigating the determinants of cardiovascular risk factors in a 1% random sample, stratified by age and gender, of the general population in nine districts of the Czech Republic. A total of 3,612 individuals aged 25–64 years were examined in 2007–2009 as previously reported.^29^ Among the 502 genotyped individuals analysed here, 106 had albuminuria. Clinpradia study (NCT01907958) was a multicenter study to evaluate the management of microalbuminuria in hypertensive patients with T2D in Canada. Its primary objective was to assess the impact of a point-of-care testing for urine albumin excretion on treatment of hypertensive T2D patients as measured by albuminuria status in primary care clinical setting. The study was performed in 2013-2014. 230 patients recruited (mean age 67 years old) in general practice clinics of Ontario and Quebec in Canada were followed for a period of 18 months. Forty percent of patients had albuminuria at study entry. The Canadian Partnership for Tomorrow’s Project (CPTP, partnershipfortomorrow.ca) brings together five Canadian regional cohorts: British Columbia Generations Project, Alberta’s Tomorrow Project, Ontario Health Study, CARTaGENE^30^ (Quebec) and the Atlantic Partnership for Tomorrow’s Health. 10,802 subjects have been genotyped up to now and 601 had T2D. 488 (mean age 58 years old) were used to calculate the AUCs for myocardial infarction and stroke after filtering for Caucasian origin, no missing data, no mismatch and others. Controls were defined as T2D individuals with no complications.

The genetic sub-study of ADVANCE was approved by 120 local ethics committees and ADVANCE participants signed a separate consent form for the genetic sub-study. DNA samples were collected only from those who signed the consent form for the genetic sub-study. The project has been approved by the CHUM Research Ethics Board since June 2014 under the OPTITHERA Program (REB #14.097) and under the Clinpradia Project since February 2016 (REB #15.335).

### Genotyping

ADVANCE participants were genotyped using the Affymetrix Genome-Wide Human SNP Arrays 5.0 or 6.0 or the Affymetrix UK BioBank Axiom arrays (Affymetrix, Santa Clara, California, USA) while participants in the Post-MONICA were genotyped with the Genome-Wide array 6.0 and those in Clinpradia with the Affymetrix UK BioBank Axiom array. Subjects of CPTP were genotyped using Affymetrix UK Biobank Axiom or GSA Illumina arrays. A quality control filtering step was applied to the genotype calls as described in our previous work^25^ in which we showed that a principal component (PC) analysis of genotyped Caucasian participants of ADVANCE using EIGENSOFT 3.0 package identified two main sub-groups of Balto-Slavic and Germano-Celtic ethnic backgrounds (Figure S1 (a)). Here, we selected a subset of 34,570 independent SNPs to perform the PC analysis that we used to adjust for ethnicity in all these cohorts (Figure S1 (b and c)).

### Creation of PRS

We selected SNPs associated to 26 risk factors or outcomes of T2D that we grouped into 9 PRS as shown in Supplementary Table S1. These 9 PRS include SNPs associated to diabetes, obesity, blood pressure, albuminuria, glomerular filtration rate (GFR), biomarkers levels, lipids, cardiovascular diseases, and low birth weight. We extracted 598 SNPs listed in Table S2 together with their effect size (β) from summary statistics of large-scale GWAS included in the National Human Genome Research Institute GWAS catalog and using HuGE navigator. As an example, 158 SNPs associated to cardiovascular complications group were derived from GWAS involving around 865,000 participants. To test the effect of these 598 SNPs in ADVANCE participants, we used the additive model, assuming that each SNP is independently associated with risk. At first, we constructed 9 weighted PRS (wPRS) for the nine risk groups, as different SNPs contribute with different weights to the PRS value. We calculated these wPRS by summing the product of the number of risk alleles for each patient by the effect size of those SNPs i.e.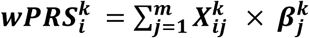, where 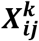 is the allele frequency of *i*^*th*^ subject in *j*^*th*^ SNP for *k*^*th*^ phenotype and β is the effect size attributed to the SNP for the same phenotype in the original GWAS (Supplementary Figures S2 and S3 and Table S2). The effect size attributed to each SNP was obtained from the same group of complications or risk factors. As an example, the effect size of a SNP associated to diabetes was used in the construction of the PRS-Diabetes only. If the same SNP was also shown to be associated to albuminuria in meta-analysis, the effect size used in the other PRS was the one from the original meta-analysis of albuminuria and not the effect size derived from the meta-analysis of T2D. Five genes (TCF7L2, ADCY5, FTO, GCKR and HNF1A) were used in two PRS. The wPRSs were computed for each of the 26 risk predictors and scaled before being additively grouped into nine PRSs of risk categories and used as variables in the logistic regression models adjusted for sex, age at diagnosis, diabetes duration, and PC1 of ethnicity as illustrated in Figure S3. Details of statistical analyses, genotyping, imputation as well as the stepwise approach for selection of SNPs and creation of PRS are included in Supplementary text and Figures S2 and S3 and Tables S1 and S2.

### Hierarchical clustering

We performed unsupervised hierarchical clustering (hclust complete method) as described by the R Core Team^31^ on the Euclidian distance matrix of the predicted risk values of our models (myocardial infarction, stroke, heart failure, major macrovascular events and cardiovascular, and all cause death). Heatmaps were constructed using R heatmap.2 from gplots library.^31,32^

### Prediction using clinical risk scores

For comparison, we calculated the ADVANCE clinical risk score that includes age at diagnosis, known duration of diabetes, sex, pulse pressure, treated hypertension, atrial fibrillation, retinopathy, HbA1c, UACR, and non-HDL cholesterol^33^ and the widely used Framingham (FRS) score that uses age, sex, total cholesterol, HDL cholesterol, smoking status, diabetes, systolic blood pressure and blood pressure treatment as predictors.^34^

## Results

### Predictive performance of PRS for individual and combined T2D complications

The baseline characteristics that include age, sex, age at diagnosis of T2D, diabetes duration, HbA1c, blood pressure, and renal function (eGFR and urinary albumin creatinine ratio (UACR)) of 4098 subjects of European descent from ADVANCE are shown in Table S3 along with 11,140 subjects of the entire set. During the median trial period of 4.9-years of ADVANCE, the 4098 genotyped participants had 334 microvascular, 559 macrovascular events and 844 combined micro-macrovascular events and 283 cardiovascular and 549 all cause deaths as detailed in Table 1.

**Table 1:** Performance of the polygenic risk model in predicting main T2D outcomes.

| Outcomes (n) | Total |  |  |  |  | High Risk (HR) |  |  | Low Risk (LR) |  |  | HR/LR |
| --- | --- | --- | --- | --- | --- | --- | --- | --- | --- | --- | --- | --- |
|  | PP (%) | AUC (95% CI) | AUC<sup>1</sup> (95% CI) | PPV | NPV | PP (%) | PPV | NPV | PP (%) | PPV | NPV |  |
| Macroalbuminuria (150) | 3.8 | **0.64**<br>(0.59-0.68) | **0.65**<br>(0.60-0.71) | 5.5 | 97.6 | 6.0 | 6.7 | 96.1 | 2.1 | 2.7 | 98.7 | 2.9 |
| New/worsen nephropathy (198) | 4.8 | **0.64**<br>(0.60-0.68) | **0.66**<br>(0.62-0.71) | 8.2 | 96.6 | 7.4 | 10.2 | 94.4 | 2.5 | 3.1 | 98.4 | 3.0 |
| New/worsen retinopathy (151) | 3.7 | **0.68**<br>(0.64-0.73) | **ND** | 6.1 | 98.2 | 6.4 | 11.4 | 95.4 | 1.7 | 2.1 | 99.0 | 3.8 |
| Major microvascular (334) | 13 | **0.67**<br>(0.64-0.70) | **0.68**<br>(0.65-0.72) | 22.4 | 91.7 | 22.2 | 32.3 | 81.8 | 6.5 | 7.4 | 94.5 | 3.4 |
| Major macrovascular (559) | 20.6 | **0.68**<br>(0.65-0.70) | **0.73**<br>(0.70-0.76) | 30.2 | 87.0 | 32.7 | 41.8 | 77.3 | 10.8 | 14.9 | 94.3 | 3.0 |
| Stroke (154) | 4.1 | **0.66**<br>(0.62-0.71) | **0.74**<br>(0.69-0.79) | 8.4 | 97.3 | 7.1 | 9.3 | 94.8 | 2.1 | 2.8 | 99.0 | 3.4 |
| Myocardial infarction (192) | 6.5 | **0.66**<br>(0.62-0.70) | **0.69**<br>(0.65-0.74) | 9.8 | 96.5 | 10.3 | 13.5 | 93.9 | 3.0 | 3.8 | 98.7 | 3.5 |
| Heart failure (225) | 5.7 | **0.68**<br>(0.65-0.72) | **0.74**<br>(0.70-0.78) | 9.9 | 96.7 | 9.9 | 16.9 | 92.7 | 2.4 | 2.7 | 100.0 | 4.2 |
| All cause death (549) | 13.4 | **0.69**<br>(0.67-0.72) | **0.74**<br>(0.72-0.77) | 22.1 | 91.9 | 22.9 | 31.2 | 84.5 | 6.3 | 9.3 | 94.6 | 3.6 |
| Cardiovascular death (283) | 7.4 | **0.72**<br>(0.69-0.75) | **0.78**<br>(0.74-0.81) | 14.8 | 96.2 | 14.6 | 20.6 | 90.0 | 2.9 | 3.5 | 98.2 | 5.0 |

The polygenic risk model is composed of the nine PRS adjusted for PC1, sex, age at diagnosis and diabetes duration. The number of cases as well as the prevalence of each outcome are indicated. AUCs and percentile-based confidence intervals (CI) were estimated from ROC curves and calculated from the predicted risks derived from the regression models. The controls used did not have the specific outcome at any time
during the study. $AUC^1$ was calculated using a control group that includes normotensive subjects. Positive predictive and negative predictive values were adjusted for the prevalence of the specific outcome. The outcome “Major macrovascular events” is a composite of non-fatal myocardial infarction, non-fatal stroke, or cardiovascular death. Microalbuminuria is defined as urinary albumin creatinine ratio of 30 to 300 mg/g at the end of the study. Macroalbuminuria is defined as urinary albumin creatinine ratio of >300 mg/g at the end of the study. New or worsening nephropathy is defined as the development of macroalbuminuria, doubling of serum creatinine to a level of at least 200 mmol/l, ESRD is defined as a need for dialysis or renal transplantation, or death due to renal disease. The outcome “Major microvascular events” is a composite of end-stage renal disease (ESRD), defined as requirement for renal-replacement therapy, death from renal disease, requirement for retinal photocoagulation or diabetes-related blindness in either eye.

The AUCs of Table 1 represent the discrimination between incident cases, defined as having an outcome during the ADVANCE trial (free of outcome at baseline) from controls that did not have a specific outcome at any time during the study. As shown in Table S4, the inclusion of 9 PRS based on related traits improved the predictive performance of the model. The AUCs of macroalbuminuria, new or worsening nephropathy, myocardial infarction, stroke or heart failure were higher with 9 PRS than with only one PRS composed of variants associated with the outcome itself (Table S4). The polygenic risk model adjusted for sex, ethnicity, age at onset and diabetes duration had AUCs ranging from 0.64 (95%CI: 0.59-0.68) for macroalbuminuria to 0.72 (95%CI: 0.69-0.75) for cardiovascular death (Table 1). The highest AUCs (*AUC*^*1*^ in Table 1) were observed for all outcomes when cases were compared to normotensive individuals who did not have a specific outcome at any time during the study reaching *AUCs*^*1*^ of 0.78 (95%CI: 0.74-0.81) for cardiovascular death, 0.74 (95% CI: 0.69-0.79) for stroke and heart failure (95%CI: 0.70-0.80) and 0.69 (0.65-0.74) for myocardial infarction (Table 1). The AUCs were not different between individuals of the whole cohort treated or not with ADVANCE intensive therapies and adding treatment assignment as a covariate in the model made no differences to the AUCs (Table S5). As expected, the AUCs of macrovascular events were lower in the quarter of individuals assigned to the combined ADVANCE therapy compared to the AUCs of its control group, as the intensive treatment reduced the differences between cases and controls (Table S6).

The AUCs of the PRS model compared to ADVANCE and Framingham clinical scores are shown in Table S7. The PRS AUCs for macrovascular events, myocardial infarction, stroke, heart failure and total and cardiovascular death were not statistically inferior from those obtained with the ADVANCE clinical score, knowing that the latter must be considered “optimistic” as our subset of patients was part of the population from which it was developed and included such clinical outcomes as atrial fibrillation, albuminuria, low eGFR and retinopathy.^35^ They were significantly higher than those obtained with the widely used Framingham risk score (Table S7).

### Risk stratification

The risk of T2D outcomes and death increased exponentially according to PRS deciles, rising sharply at the last three deciles of the distribution, suggesting that 30% is the threshold for high risk individuals in ADVANCE (Figure 1). For instance, the top three deciles of ADVANCE participants with the highest PRS had 3.1-fold increased risk of major micro- and macrovascular events (p=6.3×10^−21^ and p=9.6×10^− 31^, respectively), 4.4-fold (p=6.8×10^−33^) increased risk of cardiovascular death and 3.1-fold (p=1.9×10^−30^) higher risk of all-cause death than the remainder of participants. The threshold of 30% was confirmed by an unbiased, unsupervised hierarchical clustering analysis that identified three clusters of individuals representing 37.1%, 33.5%, and 29.4% of ADVANCE individuals having a *low, medium*, or *high* genetic risk of major macrovascular events including myocardial infarction, stroke, heart failure, and cardiovascular death (Figure 2a, left panel). The incidence of cardiovascular death was 3.8-fold higher in individuals at high (11%) vs low genetic risk (2.9%) (p=1.5×10^−13^) meaning that a total of one fifth (20%) of individuals of the high genetic risk stratum have died during the five years of the ADVANCE trial compared to only 5% in the low risk category (Figure 2b, right panel). The difference between these two groups was also highly significant for microvascular events (including increase of albuminuria and decrease of eGFR) known to contribute to the high level of mortality in high risk individuals (Figure 2b, right panel).

**Figure 1:**
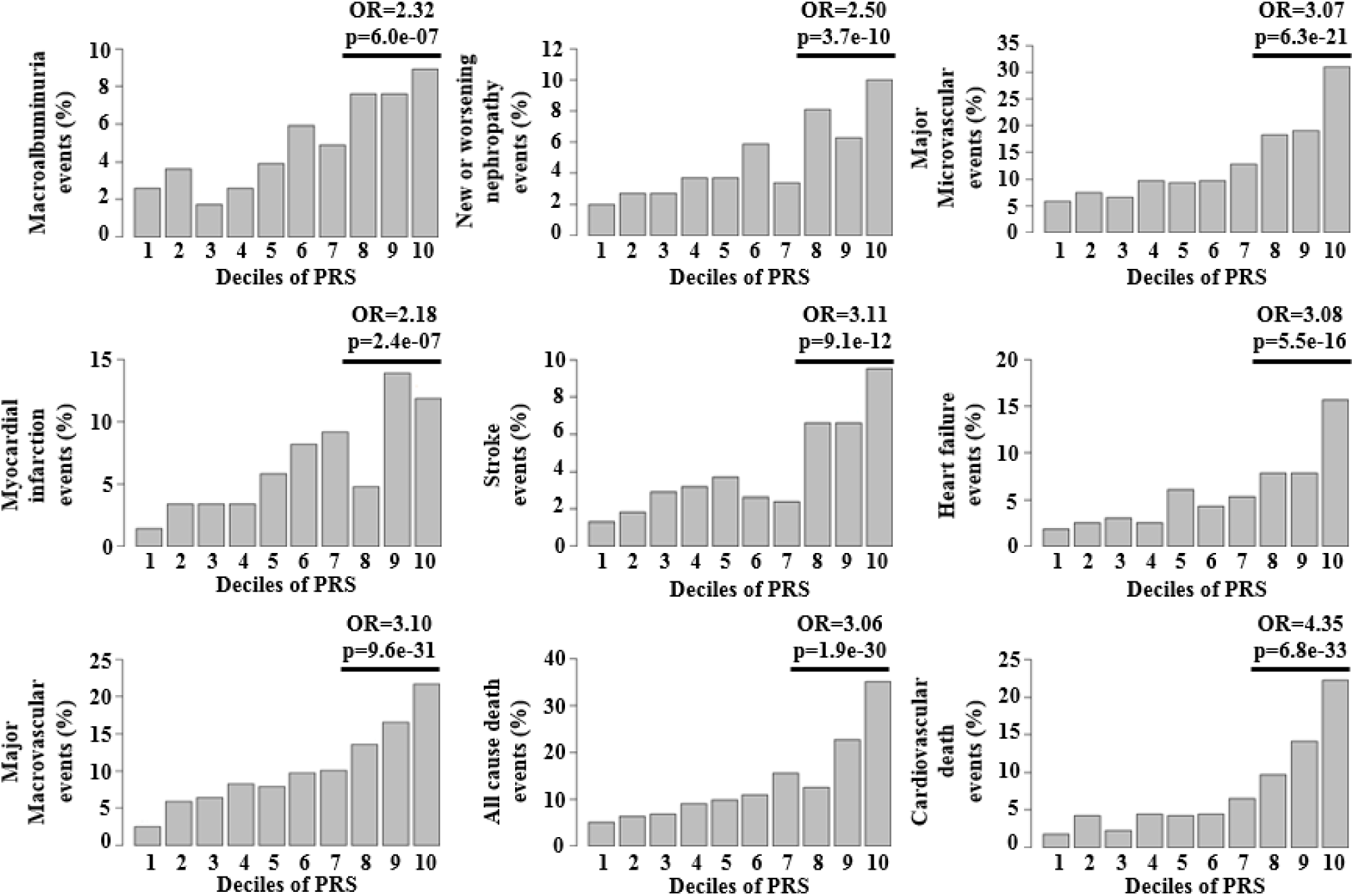
Percentage of events by deciles of PRS. OR was obtained by comparing the top 30% of distribution with the remainder of population.

**Figure 2:**
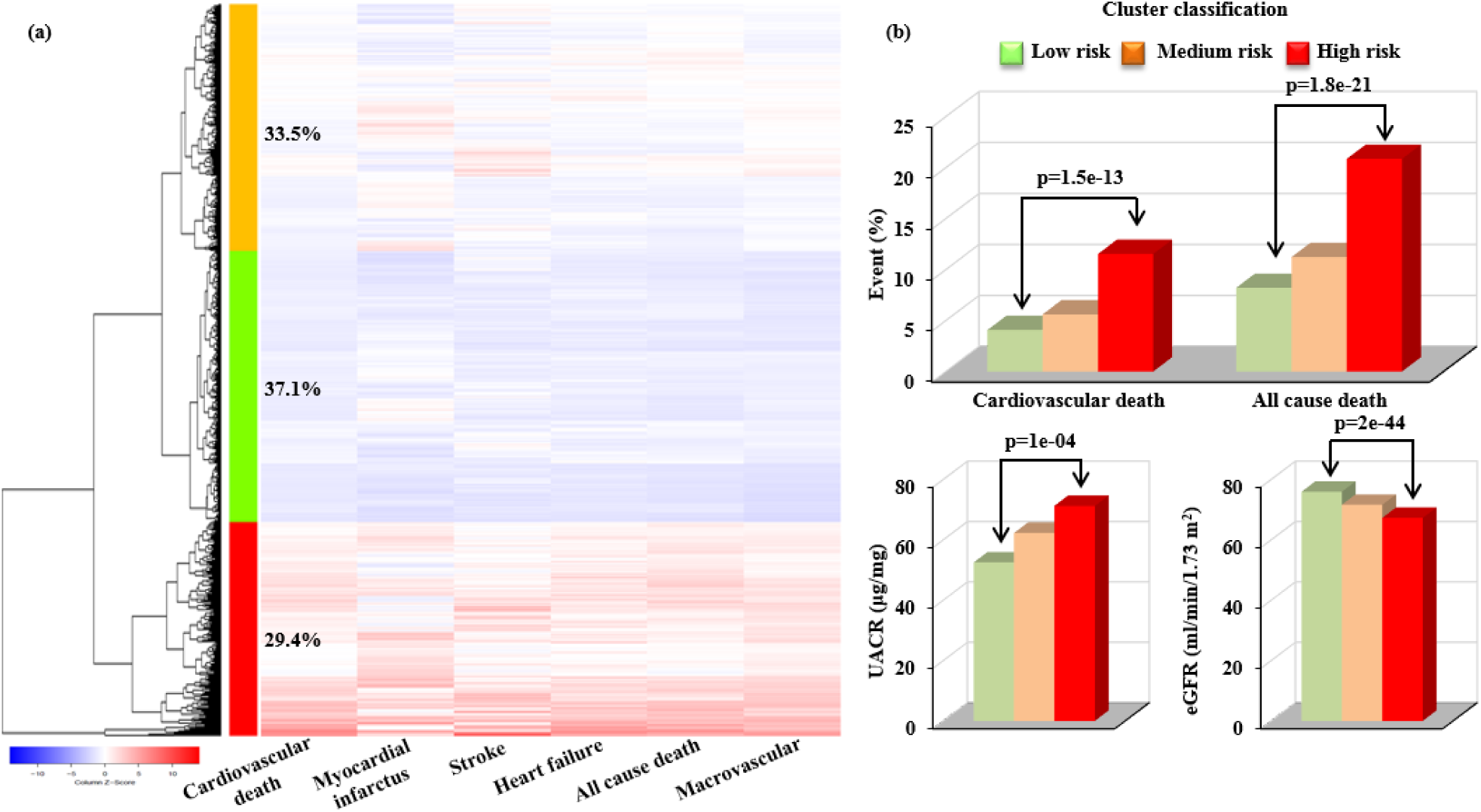
Clustering of combined macrovascular disease risk by PRS using unsupervised hierarchical clustering algorithm. This clustering method identified three clusters of individuals with *low (green), medium* (orange), or *high* (red) risk for combined macrovascular risk representing 37.1%, 33.5%, and 29.4% respectively of ADVANCE patients. a) The PRS values for each participant and each outcome were represented by Z-score (blue color: low risk score & red color: high risk score) in the heat map. b) The incidence (%) of cardiovascular and all cause death were compared between the three clusters. UACR and eGFR values were determined in the three clusters. **Abbreviation: UACR:** Urinary albumin creatinine ratio, **eGFR:** Estimated glomerular filtration rate based on CKD-EPI formula.

The negative (NPV) and positive (PPV) predictive values of the specific outcomes in ADVANCE are shown in Table 1, in the total cohort and in the high-third and low-third risk groups, separately. The NPV exceeded 95% for most outcomes in the low-risk group meaning that less than 5% of these individuals developed an outcome, suggesting low benefit of treating individuals of this group. As the incidence of events is generally low, our predictive model had inevitably lower precision, resulting in lower PPV: However, the ratio of events between the high and low-risk individuals demonstrated a clear enrichment of at-risk individuals in the high-risk group, even for less frequent events such as macroalbuminuria, retinopathy or stroke (HR/LR=2.9, 3.8 and 3.4, respectively) (Table 1).

### Contribution of the other predictors to the polygenic risk prediction model

We used individuals of European ancestry of ADVANCE as targets of our PRS as the great majority of GWAS have been performed in individuals of European descent. Furthermore, we previously reported significant differences in several of the diabetes outcomes between Europeans of Slavic and Celtic origins and it was therefore important to adjust the weights of risk alleles according to the descent, using PC1^25^. Although PC1 by itself has a variable impact on the AUC, its addition generally improved the prediction performance of the PRS of all outcomes as shown in Table S8.

Similarly, at the onset of diabetes, genomic components and information about sex and age at onset of diabetes have similar AUC values. The highest AUCs are obtained by the combination of non-genomic and genomic factors. For individuals who already have diabetes, information about diabetes duration increases the AUCs of most outcomes, particularly for such late outcomes as heart failure and death (Tableau S9).

Stratification of ADVANCE participants into equal thirds of PRS and age of onset showed that the highest risk of microvascular events was seen in carriers of high PRS with OR _High vs Low_=1.53 (1.08-2.17), p=0.017 and younger age at onset of T2D (OR _Old vs Young_=0.61 (0.43-0.87), p=0.0057) (Figure 3). This contrasts with macrovascular events, for which the highest risk was seen in the highest PRS group with OR _High vs Low_=2.78 (2.02-3.81), p=2.6×10^−10^ independently of the age of onset of diabetes. It is noteworthy that the stratification capacity of the PRS was best in people with earlier onset of T2D for both major micro- and macrovascular events, as shown by the p trend values of 4.6×10^−3^ for major microvascular and 1.7×10^−7^ for major macrovascular events (Figure 3).

**Figure 3:**
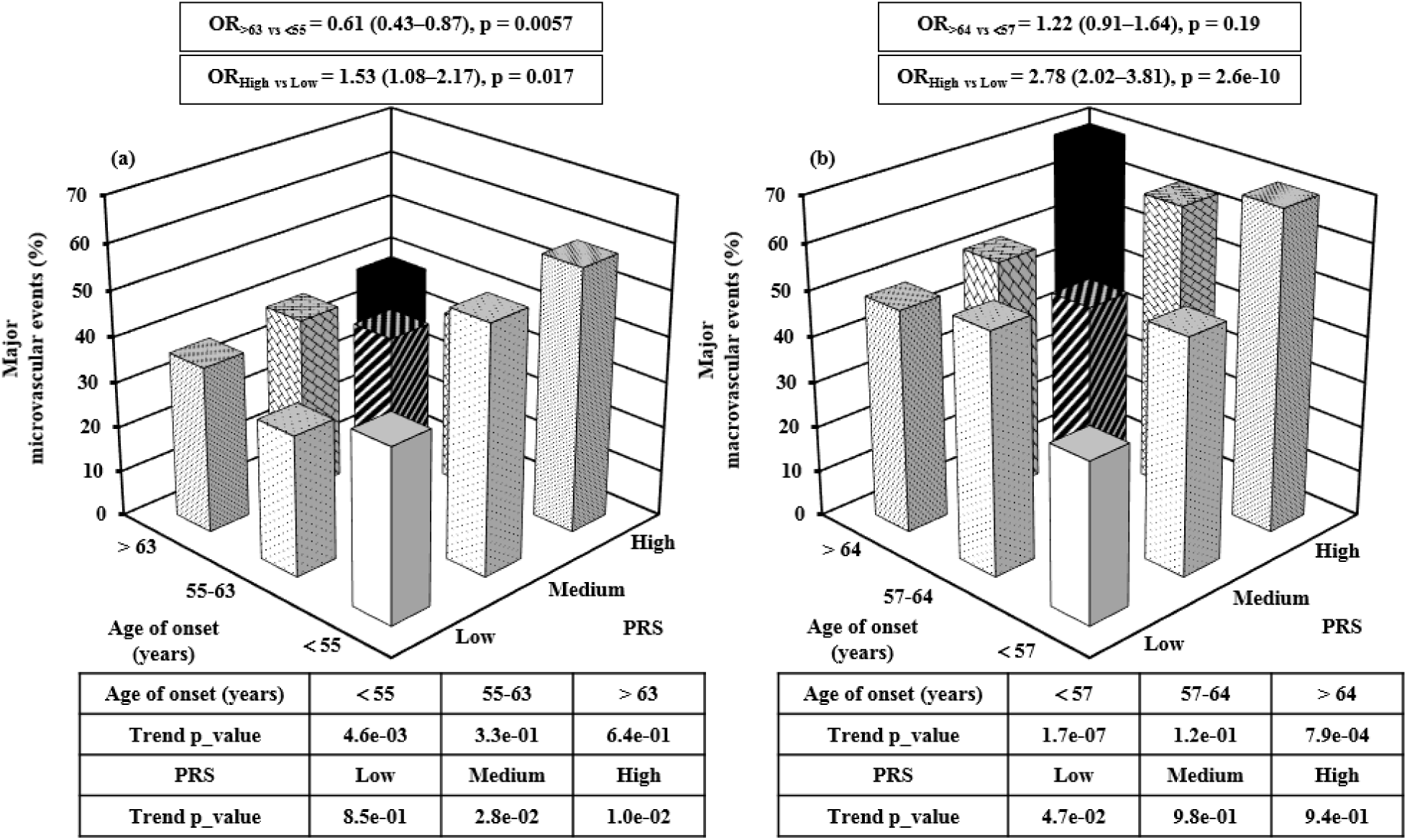
Frequency of major microvascular and macrovascular events by PRS and age at diagnosis of diabetes strata. ADVANCE participants were stratified into equal thirds of low, medium and high PRS strata and of <55, 55-63 and >63 years of age at diagnosis of diabetes. The controls used are normotensive subjects with no major microvascular (a) and major macrovascular (b) events at any time during the study. OR were calculated between low and high PRS and between age of onset <55 years old and >63 years old for microvascular and between age of onset <57 years old and >64 years old for macrovascular outcomes. The trend testing was done within formal regression analysis using parametric method separately for different age categories and PRS strata. Major macrovascular and major microvascular events are defined in Table 1.

We investigated further the contribution of the PRS and age at onset of T2D to the prediction of albuminuria and low eGFR, two independent predictors of cardiovascular and renal outcomes in T2D^33^. While age was the dominant predictor of low eGFR (Figure S4 (c)), the PRS was a major predictor of albuminuria particularly in younger diabetic patients (Figure S4 (a)) and in persons with younger age at T2D diagnosis (Figure S4 (b)).The only significant interaction was observed between the PRS and age of onset (p=0.02), such that the prediction capacity of albuminuria by the PRS was highest in people with early onset of diabetes (before 56 years old) (Figure S4 (b)).

### Calibration and replication

The polygenic risk model was well calibrated (expected vs observed event rates are similar) for cardiovascular death in the whole population (p=0.67) with better fit (closer the p value is to 1, better is the fit) for males (p=0.66) than females (p=0.48) and for Slavic (p=0.77) than Celtic (p=0.44) individuals. The best fit was observed for all cause death, p-values exceeding 0.8 in both Celtic and Slavic individuals (Figure 4). The prediction of albuminuria using the polygenic risk model was replicated in two independent population cohorts using outcomes available in each study. An AUC of 0.62 (95%CI: 0.53-0.71; n=156) was obtained in Clinpradia (Table S10). The PRS reached an AUC of 0.56 (95%CI: 0.50-0.62; n=502) in Post-MONICA individuals who had borderline diabetes and were younger (mean age 59 years old) than participants in ADVANCE (Table S10). The prediction of myocardial infarction and stroke with the PRS was assessed in the CPTP pan-Canadian population cohort. AUCs of 0.78 (95%CI: 0.68-0.89) for myocardial infarction and of 0.80 (95%CI: 0.63-0.97) for stroke were obtained in people with T2D who were younger (mean age 58 years old) than those from ADVANCE.

**Figure 4:**
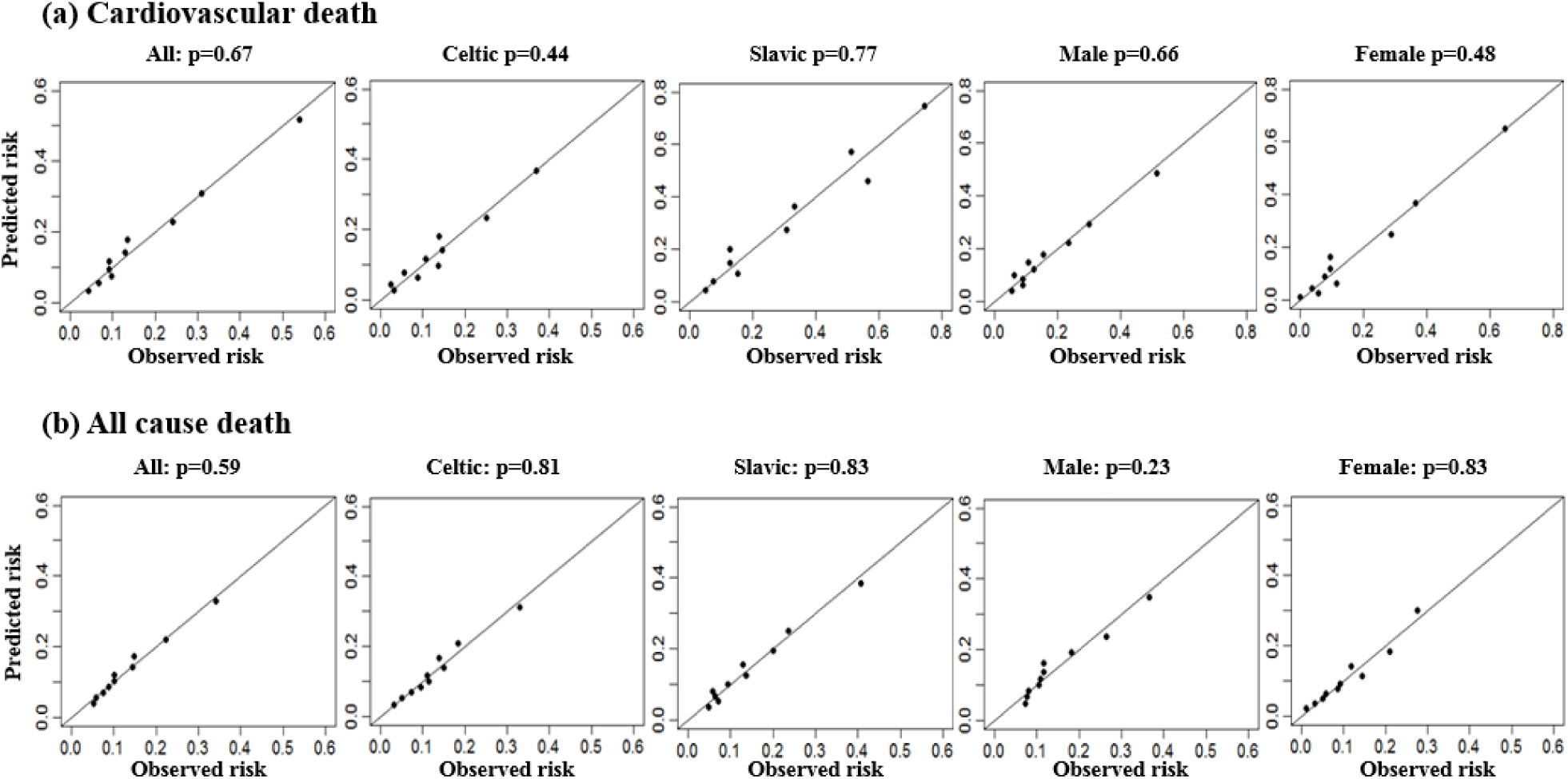
Hosmer-Lemeshow test. Ethnic- and sex-specific calibration plots for the ADVANCE cardiovascular (a) and all cause death (b). The test is used to assess the goodness of fit for logistic regression models. A value of 1 means that the expected and observed event rates in subgroups are similar. The model is therefore called well calibrated. The subgroups are sex and ethnicity.

### Clinical utility of the PRS

Even though our primary aim was not to develop a model that outperforms existing clinical scores but one that can predict as early as possible, it ought to be noted that the PRS improved that prediction of T2D outcomes of the two clinical scores, ADVANCE and Framingham. For instance, the net reclassification index (NRI) was 37% for myocardial infarction and 61% for cardiovascular death of people initially classified by the Framingham risk score and was 40% for myocardial infarction and 24% for cardiovascular death of those initially classified with the ADVANCE clinical score (Table 2). The cumulative incidence of death was significantly different (p<0.0001) between individuals with low, medium, and high PRS (Figure S5). We also noted that the response rates to the intensive therapies administered in ADVANCE were considerably higher in the high-risk compared to the low-risk groups. Intensive blood pressure control achieved during ADVANCE led to a significant reduction of total death (HR=0.797, p=0.046) and cardiovascular death (HR=0.677, p=0.009) in individuals within the highest third of PRS and these reductions remained significant during ADVANCE-ON (Figure S5, left panel). Again, in line with clinical observations, no such benefit was observed for total and cardiovascular death with intensive glycemic control (Figure S5, right panel), whilst it was observed for ESRD in individuals carrying the highest PRS values (HR=0.345, p=0.043 in ADVANCE) remaining significant at the end of ADVANCE-ON (HR=0.455, p=0.026) (Figure S6). Fifty-nine percent of ESRD cases occurred in the highest PRS third (Figure S6). The reduction of cardiovascular death by combined blood pressure and glycaemia treatments was restricted to the high PRS risk group (HR=0.612 (0.404-0.929), p=0.021) and remained significant during ADVANCE-ON (Figure 5) and the number needed to treat (NNT) to prevent one cardiovascular death could be reduced by as much as 5-fold. For instance, NNT=12 (p=0.006) in the high-risk third compared to NNT=64 (not significant) in the low-risk third.

**Table 2:** Net reclassification index adding the PRS to Framingham & ADVANCE clinical risk scores.

| Outcomes | Reclassified predicted risk with PRS (%) |  |  |  |
| --- | --- | --- | --- | --- |
|  | Framingham |  | ADVANCE |  |
|  | Net reclassified | Reclassified<br>Decreased/Increased | Net reclassified | Reclassified<br>Decreased/Increased |
| Major macrovascular | 38 | 32/52 | 20 | 14/35 |
| Myocardial infarction | 37 | 25/56 | 40 | 24/59 |
| Stroke | 37 | 26/61 | 40 | 23/58 |
| All cause death | 45 | 38/57 | 23 | 16/38 |
| Cardiovascular death | 61 | 46/72 | 24 | 16/45 |

Reclassification of predicted risk (%) with the addition of PRS to Framingham and ADVANCE scores for macrovascular outcomes of T2D. Bootstrapping was performed with 1000 iterations.

**Figure 5:**
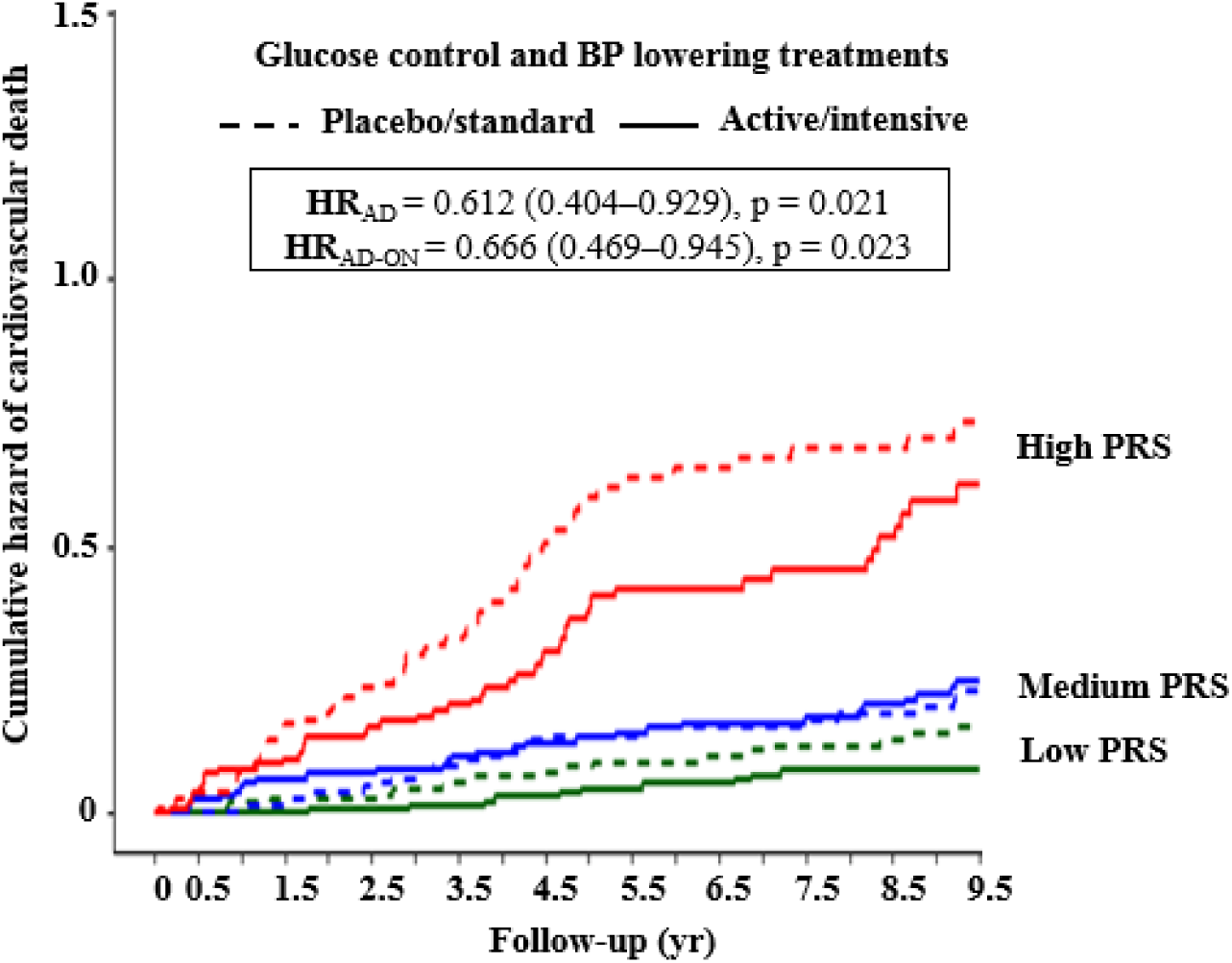
Cumulative hazard plots of cardiovascular death stratified by both glucose and BP lowering treatment and risk. Adjusted cumulative hazard curves for 9.5-year cardiovascular death by combined intensive blood pressure and glucose lowering treatment arms in the high, medium, and low PRS thirds. The controls used are normotensive subjects. The effect of BP and glucose lowering treatment was significant for individuals included in the high risk third (p=0.021 at year 4.5 end of ADVANCE trial, and p=0.023 at year 9.5 end of ADVANCE-ON follow-up).

## Discussion

Initial genetic prediction of T2D^10^, kidney diseases,^36^ stroke,^37^ or cardiovascular outcomes^38^ was unsatisfactory due to the low number of SNPs employed or use of SNPs whose associations with the disease were not validated in large meta-analyses. Novel evidence suggests that for many adult-onset common diseases, a significant degree of the heritability could be captured with a large number of common SNPs detected through genome-wide genotyping and imputation^39^ and adjustment for late penetrance using age or age of onset as covariates.^40^ It has also been recently suggested that genome-wide polygenic scores that include millions of variants may be required for common diseases to identify individuals with risk equivalent to monogenic mutations.^16^ Our approach of using a more limited but well-validated common variants associated to known predictors of diseases generates similar gradients of risk and effectiveness in identifying 30% of individuals of the highest PRS category with risk equivalent to monogenic mutations (more than 3-fold risk for major micro- and macrovascular events and for cardiovascular and total death) as genome-wide polygenic scores^16^ (Figure 1).

Diabetes is associated with both microvascular and macrovascular outcomes and the line of demarcation between their pathogenetic mechanisms is unclear^19-21^. For instance, in ADVANCE, we previously reported that increases in UACR or decreases in eGFR in patients with T2D were independent predictors of cardiovascular events and mortality^22^. More recently, we showed that combination of changes in both eGFR and UACR is a better predictor of major macrovascular events than when the two are assessed separately^23^. Other groups showed that a clinical score that captures different types of complications is more powerful in predicting mortality than a simple count of complications^41^. Furthermore, we reported a polygenic overlap between ischemic stroke and kidney function^42^ and a shared genetic architecture has been revealed between T2D and blood pressure regulation^11^. Because of their common risk factors, overlap in terms of pathogenetic mechanisms and simultaneous or consecutive involvement in end-stage multi-organ failure and mortality, we included SNPs associated to micro- and macrovascular outcomes in addition to genomic variants that are associated to their common risk factors in our PRS. Our polygenic risk model is composed of 9 PRS of related traits that improve the AUC obtained with the PRS of the outcome alone.

Re-analysis of the robust PRS developed for schizophrenia demonstrated that it contains SNPs associated to ancestry.^43^ We recently published an East-West gradient in the prevalence and incidence of T2D complications in ADVANCE participants of European descents^25^ that appears to be due in part to a significant and genetically-based earlier age of onset of T2D in patients of Slavic origin. This was instrumental in distinguishing genomic from environmentally-based determinants since the increased risk of early onset of T2D and renal damage in individuals of Slavic origin persists even when they are living in such countries as Australia and Canada. We selected 34,570 independent SNPs which in our analysis best captured the genetic distance (in Cavalli-Sforza classical sense) and thus being a measure of the genetic divergence between segments of populations from different countries of same racial denomination^44^ (Figure S1). We concur with Curtis^43^ on the importance of ethnicity in the development of PRS and propose that ancestral background when relevant to diseases needs to be added to the model to improve its predictive power as does the inclusion of sex, age, age of onset, and duration of disease exposure.

The clinical utility of a prediction model refers to its ability to prevent or ameliorate adverse health outcomes by using its results to advise clinical decision-making. Potential clinical utility of genetic risk scores emerged over last few years with the demonstration that subjects in the highest genetic risk category had the largest clinical benefit from therapy.^45^ The advantage of the polygenic prediction model that we developed for T2D complications is that it can be assessed well before the onset of diabetes and apparition of outcomes. In ADVANCE, we observed that the penetrance of outcomes differs between macro- and microvascular complications^40^ and the efficacy of stratifying subjects along a gradient of PRS was better seen in younger subjects or in patients with early onset of T2D suggesting that the usefulness of a PRS is in primary prevention before target organ damage occurs. The capacity to detect subjects with the best response to medication is one of the most important results of our study. The cumulative hazard plots using PRS stratification (Figure S5) for cardiovascular and all cause death, in ADVANCE (4.9 years for the trial) and ADVANCE-ON (for a total of 9.5 years) are concordant with our published clinical reports.^24,26^ The PRS distinguished subjects who benefit the most from intensive antihypertensive and glucose lowering therapies applied in ADVANCE. Figure 5 illustrates three components of this study: 1) Subjects classified into the low PRS category did not benefit from intensive therapies compared to patients of the higher PRS thirds; 2) Combination of glucose and blood pressure intensive therapies showed the best reduction in risk, as reported in ADVANCE^27^ and finally, 3) The highest thirds of PRS had the lowest number needed to treat with combined therapies of ADVANCE i.e. PRS risk classification is clinically effective in reducing the burden of the disease.

It is generally perceived that clinical risk scores are better than genetic ones to predict the risk of outcomes. The performance of the widely used Framingham risk score for major cardiovascular diseases and major coronary artery diseases (CAD) was poor in ADVANCE subjects,^46^ probably because they were generally older, were from many countries with different ethnic backgrounds and had characteristics different from those on which the scores were developed. The PRS developed here outperformed the Framingham score, was not significantly inferior to the ADVANCE clinical score and improved the net percentages of correctly classified persons by the two clinical risk scores.

A genetic risk score derived from 204 variants representative of all the 160 CAD loci was recently published by ACCORD, a clinical trial with a design similar to that of ADVANCE.^47^ In ACCORD, the AUC for major CAD events (defined as fatal CAD events, nonfatal MI, or unstable angina) was 0.57 with their genetic risk score adjusted for age, sex, study co-variates and principal components of population structure; an AUC much lower than 0.68 obtained with a clinical risk score based on conventional clinical predictors of CAD and including history of CAD, age, sex, the ASCVD risk estimator, and the study co-variates. Our PRS (adjusted for age at onset and diabetes duration, sex, and PC1) was 0.67 for the same outcome in ADVANCE individuals. One reason could be that we improved the AUC of CAD by integrating 9 PRS of related traits (Table S4). This is also evident from unsupervised hierarchical clustering shown in Figure 2 that illustrates a highly significant difference in microvascular events (eGFR and albuminuria) between individuals classified in the high and low risk categories of macrovascular events.

A limitation of our PRS however is that Caucasian was the only racial group represented in our genotyped set from ADVANCE as others were not immediately available due to national restrictions on genetic material but clearly a targeted effort should be made to encompass human diversity.

In conclusion, the polygenic prediction model developed here is based on common genomic variants that are present at birth and a few reliable demographic variables that are routinely collected during clinical practice without requiring the presence of any clinical manifestations or initial outcomes i.e. in the pre-symptomatic phase, suggesting that its usefulness is in primary prevention. The performance of the PRS to predict main outcomes of T2D was replicated in three independent population cohorts. The highest benefit of intensive treatment administered in ADVANCE was confined to the highest genetic risk category.

## Data Availability

to complete

## Acknowledgments

This work is supported by Genome Quebec, Canadian Institutes of Health Research (CIHR), Ministère de l’économie et de l’innovation du Québec (MEIE), Consortium québécois sur la découverte du médicament (CQDM), OPTITHERA, Servier and Canada Research Chair in Predictive Genomics to PH.

## Contributors

JT and PH contributed equally to the study design, data collection, data analyses, figures, data interpretation, literature search and writing of the manuscript and performed Clinpradia clinical study. They led the ADVANCE genetic sub-study committee which include also JC, SH, and MW. MH, CL, and RA contributed to the literature search, figures, data analyses, data interpretation and writing. FH and F-CMB developed the database and contributed to the data analyses and interpretation. RT performed the statistical analyses that were reviewed by MW. JC, MM, SH, DM, BW, NP, SZ, SC, GM, DEG, AR, LL, and MW participated in the design and execution of ADVANCE trial as members of ADVANCE Management Committee and critically reviewed and commented the manuscript. AK and RC collected the data of Post-MONICA study, contributed to the analyses and critically reviewed the manuscript. MA, VB, M-JF, MH, PA contributed the CPTP analysis and critically reviewed the manuscript. PS and CH contributed to the genetic analyses and critically reviewed the manuscript.

